# Towards intervention development to increase the uptake of COVID-19 vaccination among those at high risk: outlining evidence-based and theoretically informed future intervention content

**DOI:** 10.1101/2020.06.16.20132480

**Authors:** Lynn Williams, Allyson J. Gallant, Susan Rasmussen, Louise A. Brown Nicholls, Nicola Cogan, Karen Deakin, David Young, Paul Flowers

## Abstract

**Objectives:** Development of a vaccine against COVID-19 will be key to controlling the pandemic. We need to understand the barriers and facilitators to receiving a future COVID-19 vaccine so that we can provide recommendations for the design of interventions aimed at maximising public acceptance.

**Design:** Cross-sectional UK survey with older adults and patients with chronic respiratory disease.

**Methods:** During the UK’s early April 2020 ‘lockdown’ period, 527 participants (311 older adults, mean age = 70.4 years; 216 chronic respiratory participants, mean age = 43.8 years) completed an online questionnaire assessing willingness to receive a COVID-19 vaccine, perceptions of COVID-19, and intention to receive influenza and pneumococcal vaccinations. A free text response (n=502) examined barriers and facilitators to uptake. The Behaviour Change Wheel informed the analysis of these responses, which were coded to the Theoretical Domains Framework (TDF). Behaviour change techniques (BCTs) were identified.

**Results:** Eighty-six percent of respondents want to receive a COVID-19 vaccine. This was positively correlated with the perception that COVID-19 will persist over time, and negatively associated with perceiving the media to have over-exaggerated the risk. The majority of barriers and facilitators were mapped onto the ‘beliefs about consequences’ TDF domain, with themes relating to personal health, health consequences to others, concerns of vaccine safety, and severity of COVID-19.

**Conclusions:** Willingness to receive a COVID-19 vaccination is currently high among high-risk individuals. Mass media interventions aimed at maximising vaccine uptake should utilise the BCTs of information about health, emotional, social and environmental consequences, and salience of consequences.

**Statement of Contribution:** *What is already known on this subject?:*

- Uptake of a vaccine for COVID-19 will be vital for controlling the pandemic, but the success of this strategy relies on public acceptance of the vaccine.
- Uptake of vaccinations and public confidence in vaccines has been falling in recent years.
- Evidence suggests that 74% of the French population want to receive a COVID-19 vaccination.

*What does this study add?:*

- This study found that 86% of our sample of high-risk participants in the UK are willing to receive a future vaccine for COVID-19.
- This study showed that perceived barriers and facilitators to uptake of the COVID-19 vaccination concentrated on the ‘beliefs about consequences’ TDF domain.
- This study suggests that the content of mass media interventions to improve vaccine uptake should focus on the BCTs of information about health, emotional, social and environmental consequences, and salience of consequences. These techniques should be pitched in relation to both self and, most importantly, to others.

## Introduction

Vaccination will be vitally important in controlling the COVID-19 pandemic and bringing an end to social distancing, with researchers worldwide working to develop and test a vaccine. However, the success of this strategy relies upon public acceptance of the vaccine. At the time of writing, 115 vaccine candidates are in development and there is an indication that vaccines could be available for emergency use by early 2021 (Thanh Le et al., 2020). In recent years, vaccination rates and public confidence in vaccines has been falling (Larson et al., 2016); a pattern observed in childhood immunisations, such as measles, and in adult vaccination programmes. For example, seasonal influenza vaccination uptake, even among those who are at high risk (e.g. those with chronic illnesses), is typically less than 50%, which falls substantially below the World Health Organisation (WHO) target of 75% (Jorgensen et al., 2018). Recent findings from the Wellcome Global Monitor showed that only 59% of people in Western Europe believe that vaccines are safe (Wellcome Trust, 2019). Therefore, the current study investigated the barriers and facilitators of uptake of a future COVID-19 vaccination. We then used this information, and the framework of The Behaviour Change Wheel (BCW; Michie, Atkins & West, 2014), to make recommendations about the design of interventions aimed at maximising vaccine uptake by the public.

Vaccine hesitancy was evident during the last worldwide pandemic (H1N1 “swine flu” in 2009), with data indicating that vaccination uptake was variable. Most countries reported that less than half of the target population received the vaccine (Brien et al., 2012). The target population varied by country with some countries aiming to vaccinate the whole population and others only offering the vaccine to particular groups (e.g. health professionals, children, people with chronic disease, pregnant women). A systematic review by Bish et al. (2011) examined the factors associated with vaccination uptake during that pandemic. They found that stronger vaccination intentions and higher vaccination uptake were related to the degree of threat experienced and perceptions of vaccination as an effective coping strategy. Research also showed that the majority of the public felt that they had a low risk of acquiring H1N1 (Davis et al., 2015; Seale et al., 2010). In addition, non-uptake was related to concerns about the safety of the vaccine (Han et al., 2016) and the belief that the vaccine had not been properly tested and was rushed into circulation (Fabry et al., 2011; Seale et al., 2010).

There are key differences between the current COVID-19 pandemic and the 2009 pandemic. Most notably, the 2009 pandemic was categorised as ‘moderate’ by the WHO, and most people who were infected had mild symptoms (Wu et al., 2010). In contrast, indications are that the COVID-19 pandemic is more contagious and has a higher mortality rate than the 2009 pandemic. Relatedly, the pandemic has brought severe restrictions in travel and daily activity across the globe. Consequently, perceptions of risk related to COVID-19 are likely to be higher than for the 2009 pandemic, which may in turn increase uptake of vaccination (Brewer et al., 2007).

Early evidence relating to potential COVID-19 vaccine hesitancy comes from France. In a survey of a representative sample of the French population ten days after the nationwide lockdown was introduced, 26% of respondents said they would not want to receive a future vaccine for COVID-19 (The COCONEL Group, 2020). This was higher among low-income participants (37%), and also prevalent among people aged older than 75 years (22%) who are at high-risk for COVID-19. However, it is important to note that this study was conducted at an early stage of the pandemic. Intention to receive a future vaccine may now have increased as the pandemic has progressed, with perceptions on severity potentially increasing in line with the increasing numbers of those affected. More recently, a study reported more positive views relating to a COVID-19 vaccine among Australian adults, with only 4.9% stating they would not get the vaccine, and 9.4% stating indifference. These beliefs were associated with lower education levels and health literacy, and the belief that the threat of COVID-19 had been exaggerated (Dodd et al., 2020).

The current study aimed to identify and understand the barriers and facilitators to receiving a future COVID-19 vaccine and, using the BCW as a framework (Michie et al., 2014), provide recommendations for the design of interventions aimed at maximising uptake of the vaccine among the public. The BCW provides a useful framework for designing interventions. It is based on the synthesis of 19 existing behaviour change frameworks, and has the Capability Opportunity Motivation-Behaviour (COM-B) model at the centre (Michie, Stralen, & West, 2011). The COM-B model describes behaviour as the interaction between an individual’s capability, opportunity and motivation to engage in the behaviour with six components that drive behaviour (i.e. physical capability, psychological capability, physical opportunity, social opportunity, reflective motivation, and automatic motivation). These COM-B components are linked to intervention functions (i.e. education, persuasion, incentivisation, coercion, training, enablement, modelling, environmental restructuring, restrictions) through which an intervention can change behaviour, and seven broad policy categories (i.e. guidelines, environmental/ social planning, communication/ marketing, legislation, service provision, regulation, fiscal measures). Intervention functions are then linked to behaviour change techniques (BCTs), which are the observable, replicable and irreducible active ingredients of an intervention (Cane et al., 2012, 2015; Michie et al., 2014), with BCT groupings of goals and planning, feedback and monitoring, social support, shaping knowledge, natural consequences, comparison of behaviour, associations, repetition and substitution, comparison of outcomes, reward and threat, regulation, antecedents, identity, scheduled consequences, self-belief, and covert learning.

The present study has four key research questions (RQs):

**RQ1**: What proportion of participants are willing to receive a vaccination for COVID-19 if it becomes available, and what are the associated psychological and socio-demographic factors?

**RQ2**: What are the factors that would shape the participants’ COVID-19 vaccination behaviour, and which theoretical domains are key in shaping this behaviour?

**RQ3:** What key themes are present within the most dominant theoretical domain and what do these suggest about future intervention content?

**RQ4:** What are the relevant intervention functions and associated behaviour change techniques that we can use to provide potential evidence-based and theoretically informed future intervention content?

## Methods

Data collection took place for ten days from 1^st^ April 2020, spanning the second and third weeks of lockdown in the UK. At that time, the COVID-19 vaccination was in early development, with the first human trial of the vaccine commencing on 23^rd^ April 2020 in the UK. The sample for the present study had previously been recruited for two ongoing projects examining vaccination behaviour more broadly. Ethical approval was received from the University Ethics Committee to extend these studies by contacting participants and inviting them to answer questions relating to COVID-19. Participants completed an online questionnaire assessing their views on COVID-19. The questionnaire included a free text response in order to gather qualitative data regarding barriers and facilitators to vaccination, with responses mapped to the TDF.

### Participants and procedure

The present sample comprised 527 participants (57% female) with a mean age of 59.5 years old (*SD* = 16). This represented a convenience sample recruited to purposive criteria. Of those, 311 were older adults, and 216 had chronic respiratory disease. In order to be part of the original older adult vaccination study, participants had to be aged 65+ (i.e., the age required to be offered the annual flu vaccination free of charge) and living independently. In order to be part of the chronic respiratory disease sample, participants had to be aged 18-64 with a chronic respiratory disease (e.g. asthma or chronic obstructive pulmonary disease [COPD]). These participants were originally recruited via social media adverts, partner organisations, and University participant panels. For the current study, participants received an email inviting them to take part in this additional wave of data collection relating to COVID-19. Participants accessed the questionnaire by clicking on the Qualtrics link contained within their invitation email.

### Questionnaire

#### Perceptions of COVID-19

Seven items were used to assess perceptions of COVID-19. The items were selected from previous research on perceptions of the 2009 pandemic (Rubin et al., 2009; Williams et al., 2012) and assessed: likelihood of infection (“How likely do you think it is that you will contract coronavirus (COVID-19) over the next six months”), with response options ranging from ‘very unlikely’ (1) to ‘very likely’ (4); severity of illness (“I think if I catch coronavirus it will have major consequences for my life”), timeline for the outbreak (“In my opinion, the coronavirus outbreak is going to continue for a long time”), exaggeration of the risk (“I think the media have over-exaggerated the risks of catching coronavirus”), good information (“Overall, the information about coronavirus has been clear”), and trust in authorities (“In general, I think the authorities are acting in the public’s best interest in dealing with the coronavirus outbreak”), all of which had response options of ‘strongly agree’ (5) to ‘strongly disagree’ (1); and, finally, worry (“How worried or anxious are you about coronavirus”) with response options of ‘not at all worried’ (1) to ‘very worried’ (4).

#### COVID-19 vaccination intention

We asked “If a vaccine for coronavirus becomes available, would you want to receive it?” and provided response options from ‘I definitely would not want to receive it’ (1) to ‘I definitely would want to receive it’ (5). In addition, participants were asked, “What are the factors that would influence this decision” and provided a free text response option.

#### Influence of COVID-19 on future vaccination behaviour

Participants were asked “Do you think the current coronavirus (COVID-19) pandemic will influence your decision about whether or not to receive the annual flu vaccination in the future?” with response options including ‘Yes, it will make me more likely to get the annual flu vaccine’, ‘Yes, it will make me less likely to get the annual flu vaccine’, and ‘No, it will not influence my decision’. The older adult participants were also asked this question in relation to the pneumococcal (pneumonia) vaccination.

### Analysis

For RQ1, descriptive statistics were computed, showing the proportion of participants who are willing to receive a COVID-19 vaccine, and those who think COVID-19 will influence their vaccination behaviour in general. T-tests and chi-square were then used to examine any differences in vaccine willingness across gender or deprivation level, respectively. Next, correlation analyses were performed to examine the associations between vaccine willingness, age, and perceptions of COVID-19. Missing data was minimal, typically less than 1% for most questions.

#### Content analysis of free text response

For RQ2, we performed directed content analysis of the responses based on the TDF (Atkins et al., 2017; Hsieh & Shannon, 2005). The TDF comprises 14 broad theoretical domains (i.e. knowledge; skills; social/ professional role and identity; beliefs about capabilities; optimism; beliefs about consequences; reinforcement; intentions; goals; memory, attention and decision processes; environmental context and resources; social influences; emotion; behavioural regulation), synthesising many diverse yet related theoretical constructs. It is a useful tool that can be used to describe the causal mechanisms that shape behaviours of interest. Our free text response invited people to detail their perceptions of the factors that would shape their COVID-19 vaccination behaviour. Responses were initially coded by AG against the TDF domains. LW and PF then checked the coding for a random sample of 30% of the comments. Following rounds of discussion concerning the very few areas of disagreement, 100% agreement in coding was achieved. In this way, we derived a clear sense of which theoretical domains were important in shaping COVID-19 vaccination behaviour, and which might present useful causal mechanisms to address in future interventions.

Next for RQ3, in an attempt to provide a further more granular level of suggestions for future intervention content, we conducted further thematic analysis of the most dominant theoretical domain we found within the data (Braun & Clarke, 2006). This was conducted by AG, and the themes were refined through discussion with PF. As the majority of comments (90%) were coded under the beliefs about consequences domain, thematic analysis was applied within this domain only.

Finally for RQ4, to suggest future intervention content that was both evidence-based and theoretically informed analysis we used the BCW approach. Subsequently, we mapped the relevant intervention functions and associated BCTs to the key TDF domains we had identified were prevalent within our sample (Cane et al., 2015; Michie et al., 2013). This work was conducted by LW and PF.

## Results

### RQ1 - COVID-19 vaccination willingness

Our sample characteristics (Table 1) show that this is an older sample (due to the inclusion of adults aged 65+ as one of the high-risk groups), and that the participants are generally well-educated with more than half the sample having being educated at degree or postgraduate level. The sample is well balanced in terms of gender profile (56.7% female) and deprivation category. We found that 58% of the sample (n=307) would definitely want to receive a vaccine for COVID-19 once it becomes available, and 27% (n=143) probably would want to receive it (see Table 1). However, 7% were unsure (n=38), 2% (n=9) would probably not want to receive it, and 6% (n=29) would definitely not want to receive it. There were no differences between the older adult group and the chronic respiratory group in terms of willingness to receive the vaccine, *t* (1, 524) = .279, *p*=.78, or any differences between males and females, *t* (1, 523) = 1.45, *p*=.14. There were also no differences in willingness to have the vaccine based on socio-economic status (assessed by post-code indicated deprivation quintile; *X*^*2*^ (5, *n* = 497) = 6.32, *p* = .98).

**Table 1.**
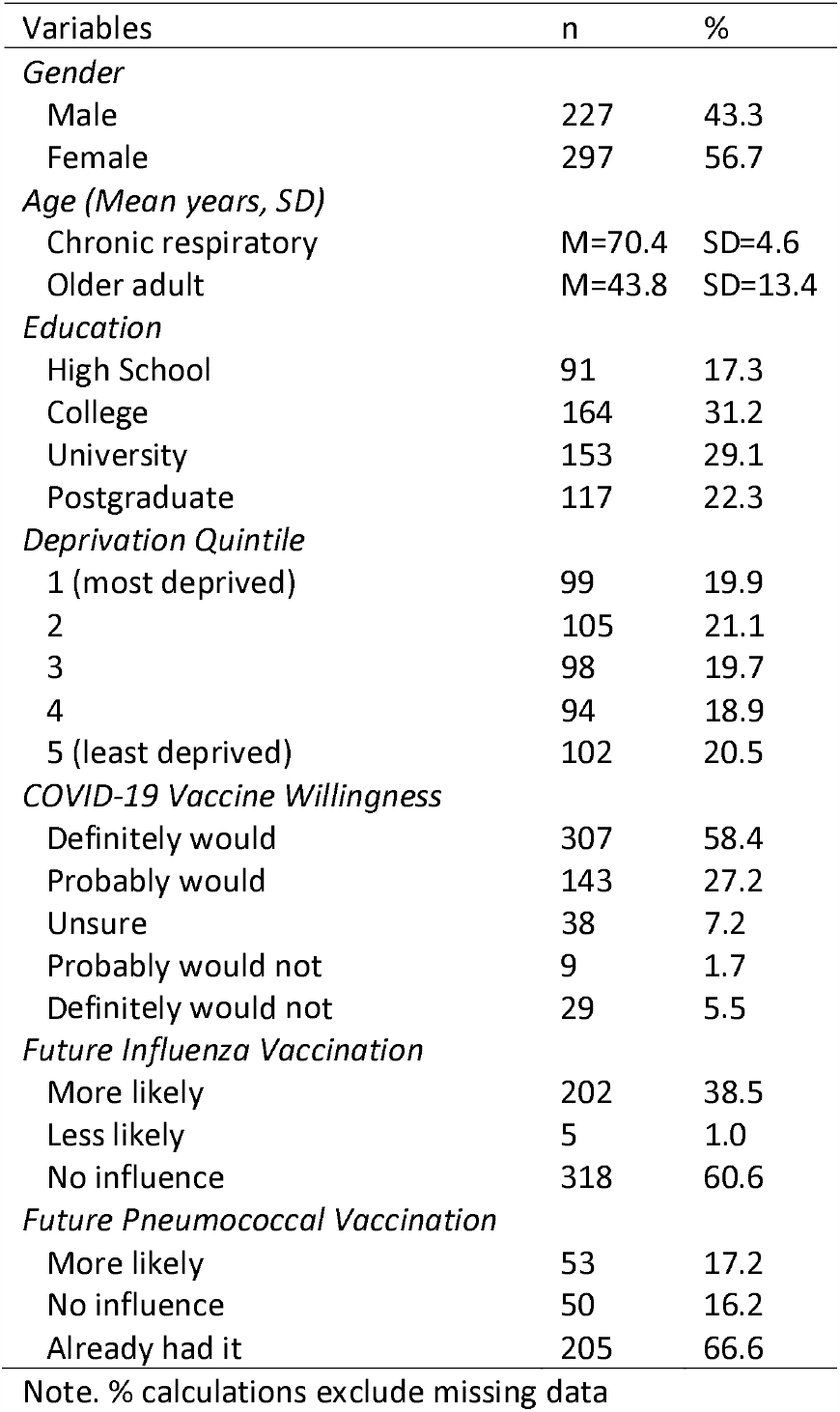
Participant demographics and vaccination intention

| Variables | n | % |
| --- | --- | --- |
| *Gender* |  |  |
| Male | 227 | 43.3 |
| Female | 297 | 56.7 |
| *Age (Mean years, SD)* |  |  |
| Chronic respiratory | M=70.4 | SD=4.6 |
| Older adult | M=43.8 | SD=13.4 |
| *Education* |  |  |
| High School | 91 | 17.3 |
| College | 164 | 31.2 |
| University | 153 | 29.1 |
| Postgraduate | 117 | 22.3 |
| *Deprivation Quintile* |  |  |
| 1 (most deprived) | 99 | 19.9 |
| 2 | 105 | 21.1 |
| 3 | 98 | 19.7 |
| 4 | 94 | 18.9 |
| 5 (least deprived) | 102 | 20.5 |
| *COVID-19 Vaccine Willingness* |  |  |
| Definitely would | 307 | 58.4 |
| Probably would | 143 | 27.2 |
| Unsure | 38 | 7.2 |
| Probably would not | 9 | 1.7 |
| Definitely would not | 29 | 5.5 |
| *Future Influenza Vaccination* |  |  |
| More likely | 202 | 38.5 |
| Less likely | 5 | 1.0 |
| No influence | 318 | 60.6 |
| *Future Pneumococcal Vaccination* |  |  |
| More likely | 53 | 17.2 |
| No influence | 50 | 16.2 |
| Already had it | 205 | 66.6 |

Note. % calculations exclude missing data

In terms of the impact of COVID-19 on future vaccination behaviour in general, 202 participants (38%) stated that COVID-19 will make them more likely to receive the annual flu vaccination in the future, with 1% (n=5) stating it would make them less likely, and 61% (n=318) saying it would not influence their decision. Among older adult participants eligible for the one-off pneumococcal vaccine, but who had not opted to receive it previously, 51% (n=53) said they would now be more likely to get the vaccine in future.

### Correlation analysis

The Pearson’s correlation analysis (Table 2) showed that willingness to receive a COVID-19 vaccination was positively associated with the belief that the COVID-19 outbreak is going to continue for a long time, and negatively associated with the belief that the media has over-exaggerated the risks of catching COVID-19. There were no significant correlations between intention to vaccinate and the other questions tapping perceptions of COVID-19, or with age. Higher levels of worry about COVID-19 were positively associated with perceived likelihood of infection, severity, and timeline, and negatively associated with media over-exaggeration and age.

**Table 2.** Correlations, means, SDs and min-max values for age, COVID-19 vaccine willingness, and perceptions.

|  | 1 | 2 | 3 | 4 | 5 | 6 | 7 | 8 | 9 |
| --- | --- | --- | --- | --- | --- | --- | --- | --- | --- |
| 1. Vaccine willingness | — |  |  |  |  |  |  |  |  |
| 2. Age | -.002 | — |  |  |  |  |  |  |  |
| 3. COVID-19 Likelihood | .051 | -.144* | — |  |  |  |  |  |  |
| 4. COVID-19 Severity | .069 | .006 | -.008 | — |  |  |  |  |  |
| 5. COVID-19 Timeline | .090* | -.017 | .178** | .280** | — |  |  |  |  |
| 6. Exaggeration of risk | -.122* | -.078 | -.132** | -.216** | -.316** | — |  |  |  |
| 7. Good information | -.006 | .205** | -.014 | .073 | -.015 | .004 | — |  |  |
| 8. Trust in authorities | .007 | .044 | .017 | .082 | .035 | -.007 | .423** | — |  |
| 9. Worry | .083 | -.129** | .224** | .364** | .288** | -.318** | -.020 | -.018 | — |
| Mean | 4.31 | 59.55 | 2.50 | 3.78 | 4.08 | 2.36 | 3.45 | 3.71 | 2.91 |
| SD | 1.06 | 16.05 | 0.64 | 0.98 | 0.74 | 1.12 | 1.00 | 1.02 | 0.82 |
| Min-Max | 1 - 5 | 18 - 92 | 1 - 4 | 1 - 5 | 1 - 5 | 1 - 5 | 1 - 5 | 1 - 5 | 1 - 4 |

Note. \* $p < .05$ , \*\* $p < .01$

### RQ2 - Content analysis of free text using the TDF

Ninety three percent of the sample provided responses to the free text question (“What are the factors that would influence this decision” [receiving a vaccine for corona virus]) resulting in 502 comments for TDF coding. Comments were typically short. Nine responses were mapped to two TDF domains as they described multiple influences on vaccination behaviour. All other responses were mapped to one TDF domain. Eight of the fourteen domains were identified across the responses: beliefs about consequences (n=455 responses), knowledge (n=27), environmental context and resources (n=11), social influences (n=7), emotion (n= 6), social/professional role and identity (n=2), optimism (n=2), and behavioural regulation (n=1). As 90% of comments were coded under the beliefs about consequences domain, thematic analysis was applied within this domain to gain more insight into the barriers and facilitators to COVID-19 vaccine uptake. A summary of the key TDF domains mapped to relevant intervention functions and BCTs is shown in Table 3.

**Table 3.**
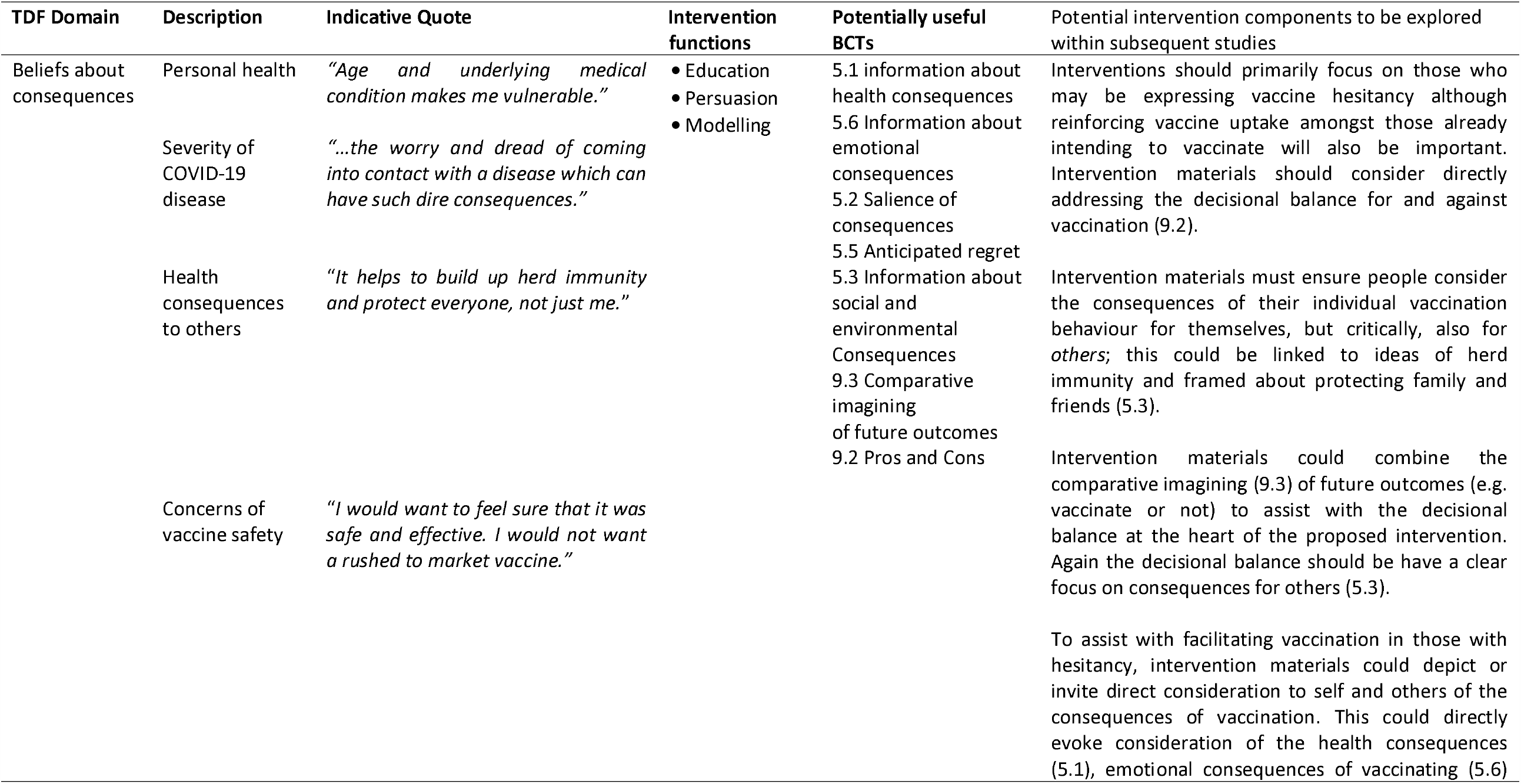
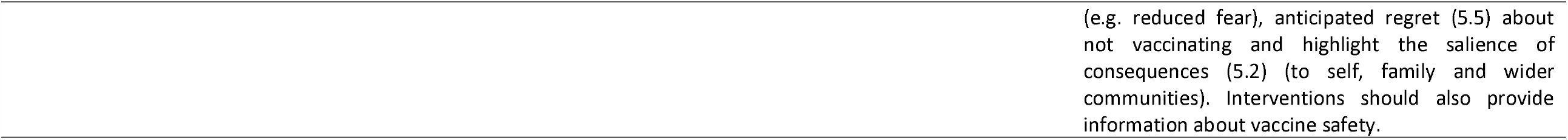
Summary of key domain mapped to intervention functions and individual BCTs.

### RQ3 - Thematic analysis of the TDF domain ‘Beliefs about consequences’

Three facilitators and one barrier were identified from thematic analysis of comments within the beliefs about consequences domain. ‘Personal health’ (n=176 responses), ‘severity of COVID-19 disease’ (n=85), and ‘health consequences to others’ (n=36), and were viewed as factors which facilitated vaccination, while ‘concerns about vaccine safety’ (n=158) was considered a barrier to vaccine uptake.

#### Personal health (facilitator)

Participants primarily described feeling particularly susceptible to contracting the virus. Risk factors included older age (“age and underlying medical condition makes me vulnerable” [female, aged 70]), having a chronic lung condition or other co-morbidities (“I have asthma so any chest infections put me at risk” [female, aged 57]), and working in a high-risk profession. Feeling vulnerable due to these risk factors, participants emphasised that vaccination against COVID-19 would provide a sense of protection (“I have a number of co-morbidities so feel it is important to take the protection which is offered” [female, aged 68]), and could help maintain their long-term health by gaining antibodies and immunity to the disease.

#### Severity of COVID-19 disease (facilitator)

Concerns of contracting COVID-19 disease and the highly contagious nature of the virus were highlighted by respondents as factors to vaccinate. The severity of contracting COVID-19, and the fear of possibly dying from the disease, were motivators for participants to vaccinate (“Having seen young healthy people pass away from this disease I don’t think I would stand a chance against it” [male, aged 36]; “…the worry and dread of coming into contact with a disease which can have such dire consequences” [female, aged 68]). Additionally, participants noted that vaccination could offer immediate protection from the disease in addition to potential outbreak ‘waves’ in the future (“this disease could come back and if we have vaccine that can prevent it then I would want it” [male, aged 70]).

#### Health consequences to others (facilitator)

Achieving herd immunity and protecting the health of others were considered benefits to vaccinating by participants. Contributing to achieving herd immunity against COVID-19 was a driving factor to vaccination for participants, with some participants considering it a ‘civic duty’ to vaccinate (“it helps to build up herd immunity and protect everyone, not just me” [female, aged 67]). Protecting others, at community and global levels, was a common factor associated with vaccine uptake, in addition to protecting family members and friends (“need to protect the human race from this new biological nightmare” [male, aged 67]). Protecting family members was especially important to participants if they felt their family members were in a high risk or vulnerable group for contracting COVID-19 (“wife has asthma so want to do it to keep her safe” [male, aged 66]). Participants also described how they would vaccinate if it assisted with ending the COVID-19 pandemic, and eliminating the need for the physical distancing and self-isolation safety measures in place (“so we never have to go through this again” [female, aged 75]).

#### Concerns of vaccine safety (barrier)

As the COVID-19 vaccination is still under development at the time of this study, barriers to vaccine uptake from participants primarily centred on the newness of the vaccine and its safety and effectiveness (“a bit sceptical as it would be a new vaccine” [female, aged 41]). Participants felt that the development of COVID-19 vaccines may be rushed, and that vaccination safety measures could be overlooked in the development process (“I would want to feel sure that it was safe and effective. I would not want a rushed to market vaccine” [female, aged 65]). Participants wanted assurance the vaccine was safe and effective at preventing disease prior to vaccination. The level of these safety and effectiveness concerns varied, with some participants stating they would want to see how safe it was before getting vaccinated (“as long as it is proven safe and effective, I would have it” [male, aged 74]), while others said they may wait years before getting vaccinated to see the longer-term effects of the vaccine (“how safe the vaccine is. I would probably wait a year or two to see how safe it is before getting it myself” [female, aged 36]). Possible vaccine side effects and severity of side effects were also cited as influences on the decision to vaccinate against COVID-19 (“how well tested the vaccine was, the side effects and interactions weighed against the chance of contracting and potential consequences coronavirus” [female, aged 60]).

Although there were many comments surrounding safety concerns of a COVID-19 vaccine, some participants felt positively about the vaccine. These participants believed in the personal and community benefits of vaccinations in general, and with reassurance of the new vaccine’s safety, would ‘certainly’ want to receive it (“trialling and testing, any potential effects a new vaccine could have. Should it be trialled and proven safe, I would take it” [female, aged 35]). Reasons to vaccinate included to prevent contraction and transmission of the virus and to lessen potential symptoms of the disease if they were still to contract the virus (“in order to reduce the chance of catching COVID-19 or at least having a milder attack than otherwise” [female, aged 71]).

### RQ4 – Potential evidence-based and theoretically informed future intervention content

Table 3 shows how our BCW analysis on the thematic analysis of ‘beliefs about consequences’ concerning vaccination behaviour. We tabularise the key thematic findings, the relevant intervention functions and appropriate behaviour change techniques. In the right hand column we operationalise this suggested intervention content into potential intervention ideas that could form the basis of further focussed intervention development work.

Overall our suggestions here focus on suggesting ways of encouraging a decisional balance amongst those with some hesitancy to enhance vaccination uptake. This could be achieved through intervention materials fostering the appraisal of an array of consequences of vaccination; for both self but particularly in relation to others.

## Discussion

To our knowledge, this study is the first in the world to examine the public’s perceived barriers and facilitators around future COVID-19 vaccination. It presents novel findings that may shape future policy, practice and intervention development. The study aimed to identify the facilitators and barriers to uptake of a future COVID-19 vaccine, specifically within high-risk populations (older adults aged 65+, and young/middle-aged adults with chronic respiratory disease). We have identified that willingness to receive a vaccination for COVID-19 is currently high among at-risk adults in the UK, with 86% indicating they would want to receive it. Given the general decline in vaccination uptake and trust observed in recent years (Larson et al., 2016), the figure of 86% is very promising. However, even in this high-risk sample, there is still a sizable proportion of people who are either undecided or who do not want to receive a COVID-19 vaccination. Reluctance to receive the vaccine was associated with the belief that the media have over-exaggerated the risks of COVID-19 and that the timeline for the outbreak will be short.

Promisingly, our data also suggest that COVID-19 may have a substantial and positive impact on vaccination behaviour in general, with 38% saying it will make them more likely to get the annual flu vaccination, and 51% saying they will now be more likely to receive the one-off pneumococcal vaccination. These figures suggest positive unintended consequences of COVID-19 on vaccine hesitancy in general. Unintended consequences are usually linked to interventions, whereby the intervention can have either positive or negative effects which were not planned by those implementing them (Oliver et al., 2019). In the current context, lockdown can be theorised as a complex public health intervention, which has had the unintended positive consequence of driving up demand for vaccines in general, as individuals seek more protection for their health. It will also be of interest to examine if there are other potentially related unintended positive consequences, such as increased uptake of screening opportunities.

Analysis of the open-ended responses showed that the beliefs about consequences TDF domain provides the greatest insight into the determinants of COVID-19 vaccination intention. We identified four key themes that chime with core components of many theories and models from health psychology that aim to explain behaviour (e.g. Rosenstock, 1974). First, personal health, chiming with the theoretical construct of personal susceptibility, which described participants feeling at risk of contracting the virus, and acted as a facilitator to vaccination as the vaccine was seen to offer protection. The second facilitator to vaccination was perceptions of the severity of COVID-19 disease, with participants noting the contagious nature of the illness and their fear of dying from it. These perceptions regarding the contagiousness and seriousness of COVID-19 are in contrast to the perceptions of the 2009 pandemic, where the public perceived that they had a low risk of acquiring H1N1 (Seale et al., 2010). The final facilitator was health consequences to others, with participants describing the importance to achieving herd immunity and protecting their loved ones. The key barrier to vaccination was concerns of vaccine safety, echoing ideas of response efficacy (e.g. Rogers, 1975), with participants noting concerns about the vaccine being rushed into development. This concern regarding the safety of a future vaccine is similar to the findings from the 2009 pandemic where non-uptake was associated with the belief that the vaccine had not been properly tested and was rushed into circulation (Fabry et al., 2011).

Using the BCW (Michie et al., 2014), we identified several intervention functions. In the current context, the functions of education and persuasion are likely to be the most useful. Education can improve knowledge of susceptibility and severity of COVID-19 and the effectiveness of vaccination, while persuasion can be used to change beliefs and encourage action towards vaccination. In terms of content of these mass media interventions, we identified a number of potential BCTs that reflected the beliefs about consequences domain of the TDF (see Table 3 for detail). As the vaccination is likely to be needed at a population-level, the mode of delivery for an intervention could be a combination of mass media (e.g.TV and radio, print media), the social media, and working closely with broadcasters and journalists to manage consistent messaging and challenge misinformation (Davis et al., 2020). A coherent media presence would enable the communication of strong descriptive and injunctive social norms concerning COVID-19 vaccination.

Further intervention development work could examine how interventions to enhance COVID-19 vaccine uptake (and uptake of the seasonal influenza vaccine this year) could potentially build upon the publics’ prior investment in COVID related behaviour change. This might include framing the intervention as associated with compliance with the lockdown, and draw explicitly upon existing intervention messaging, for example ‘*We’re in this togethe*r’ and the protective discourse implied within phrases like ‘*shielding*’. In this way, vaccination behaviours could ‘piggy-back’ onto the wider behavioural system of COVID-related change. Intervention content could connect disparate COVID-related behaviours, such as hand hygiene, social distancing and volunteering with vaccination compliance.

## Strengths and Limitations

Strengths of the study include the focus on two key populations (older adults and those with chronic respiratory disease) who are at high-risk for COVID-19. In addition, the timing of the study means that we can provide future-facing recommendations for future interventions aimed at maximising uptake of the vaccine. There are also some limitations to the present study. The duration of data collection was purposely focused during the UK peak and therefore relatively short. Our data therefore provide an indication of vaccine willingness at a particular stage of the pandemic. Further in-depth data collection is needed as the pandemic progresses in order to examine any changes in vaccination intention. In addition, as data collection was performed online this may have limited participation from older adults who do not use the internet. Moreover, our findings relate to a vaccine that is currently in development, so side effects and safety are not yet known. We also do not know if our findings would generalise to members of the public at lower risk of COVID-19, or if the presence of multiple risk factors (e.g. older age and the presence of a chronic health condition) would drive up willingness to receive the vaccine further. Furthermore, our BCW analysis is based on limited qualitative data with no quantitative element. Further in-depth research focussed on intervention development is therefore required.

## Conclusions

Intention to receive a vaccine for COVID-19 is currently high (86%) among high-risk members of the public. In addition, COVID-19 may have a positive effect on the uptake of vaccinations for annual influenza and pneumonia. Perceived barriers and facilitators to uptake of the COVID-19 vaccination concentrated on the beliefs about consequences TDF domain. Facilitators were perceptions of risk to personal health, severity of COVID-19, and health consequences to others. Concerns about vaccine safety acted as a barrier. The content of media interventions should focus on the BCTs of information about health, emotional, social and environmental consequences, as well as the salience of those consequences. This will provide the public with information about the beneficial consequences of vaccination.

## Data Availability

The data are not publicly available due to privacy or ethical restrictions.

## Acknowledgements

The University of Strathclyde supported the COVID-19 phase of data collection upon which the current paper is based. The funding for the initial phase of this research on general vaccination behaviour came from the Chief Scientist Office in Scotland (REF: HIPS/18/37 and CGA/19/52).

